# Analysis of Effectiveness of Quarantine Measures in Controlling COVID-19

**DOI:** 10.1101/2020.04.21.20074245

**Authors:** Garima Kaushik, Shaney Mantri, Shrishti Kaushik, Dhananjay Kalbande, B. N. Chaudhari

## Abstract

COVID-19 has created an interesting discourse among the people of the world particularly regarding preventive measures of infectious diseases. In this paper, the authors forecast the spread of the Coronavirus outbreak and study how the reduction of transmission rates influences its decline. The paper makes use of the SIR (Susceptible Infected Recovered) Model which is a deterministic model used in the field of epidemiology-based on differential equations derived from sections of the population. The Basic Reproduction Number (R_*o*_) represents the criticality of the epidemic in numeric terms. Forecasting an epidemic provides insights about the geographic spreading of the disease and the case incidences required to better inform intervention strategists about situations that may occur during the outbreak. Through this research paper, the authors wish to provide an insight into the impact of control measures on the pandemic. By drawing a comparison of three countries and their quarantine measures, observations on the decline of the outbreak are made. Authors intend to guide the intervention strategies of under-resourced countries like India and aid in the overall containment of the outbreak.

## 1 Introduction

Infectious disease outbreaks result in huge losses. The 1918 Spanish Flu caused around 20,000,000 deaths [1]. According to [2], as many as 3.3 billion people in the world are at the danger of contracting malaria, and a malaria related death occurs every 60 seconds. Tuberculosis has become the deadliest disease as its death rate crossed that of AIDS. In South Africa, about 80% of the people suffer from latent tuberculosis with 450,000 confirmed cases of tuberculosis in 2013 [3, 4]. At the time of this research, 7,553,182 people had been confirmed to be infected by COVID-19 and 423,349 people have died [5]. United States of America accounts for 2,010,391 confirmed and 113,7 death cases. In this paper, estimation of COVID-19 cases is done for Iran and India. Followed by analysis of intervention measures enforced at different stages in China, Italy Iran and India. The insights drawn aim to identify the best possible approach to control the pandemic of COVID-19. [6] presents the clinical qualities of the infection by analyzing 1099 patients across 552 hospitals. Of the 1099, 43.8% presented with fever whereas 88.7% developed this symptom during hospitalization and 67.8% had a cough. Only 3.8% presented with Diarrhea. The paper also defined the mean incubation period at 4 days. Of the 1099 patients, 926 patients were classified non severe and 173 patients were classified severe, on admission. The severe category of patients belonged to an older age group and had a higher prevalence of underlying illnesses as compared to non-severe patients. According to research on inanimate surfaces, human corona viruses have the capability to be infectious for up to 9 days at room temperature, however, at temperatures greater than or equal to 30 C, they are in-fectious for a shorter period [7]. Research on the effect and feasibility of containment measures has been carried out for the H1N1 outbreak in [8]and in [9]. In [9] the paper assumed that isolation could prevent transmission completely. Up to a 1000 simulations were run by considering varying values of Ro, total cases that existed initially etcetera. It considered two factors: infections resulting from a single individual and transmission that occurred before symptoms appeared. The research agreed that it is crucial to quickly follow the onset of symptoms by isolation. The isolation delay factor, as well as the number of cases to begin with, had a major influence in containing the outbreak. Sub-clinical infections might go unreported and cause a surge in transmissions. The SIR Model which was introduced for modeling influenza came forth in the early 1900s [10]. [8] describes an extension of the SIR model with three new behavioral interventions- Q_*s*_, Q_*e*_, and Q_*i*_. Where Q_*s*_ represents susceptible people that can be quarantined, Q_*e*_ is asymptomatic and not yet infectious individuals that are quarantined and Q_*i*_ are infected individuals that can be quarantined once determined as such. Discussions on how behavioral interventions that involve social distancing measures such as the closure of schools, quarantine or travel-based restrictions reduced the number of cases of influenza in [11], [12], [13], [14], [15], [16]. Some papers evaluate a combination of behavioral and biomedical interventions and determine them extremely effective [10, 12, 15, 16]. There is agreement among all papers and how implementing such strategies early on in the epidemic is the crucial part.

## 2 Materials and Methods

### 2.1 The SIR Model

The paper makes use of the SIR Model to forecast the progress of COVID-19. An SIR Model is a mathematical and an epidemiological model which generates the number of infections theoretically during a certain time period. The model derives its name from the coupled equations related to the susceptible population *S*(*t*), the population infected *I*(*t*), ad the population that has recovered *R*(*t*). An individual who is susceptible can become infected. An infected individual may recover or die. But an infected individual cannot become susceptible again as in Figure 1.

**Fig. 1.**
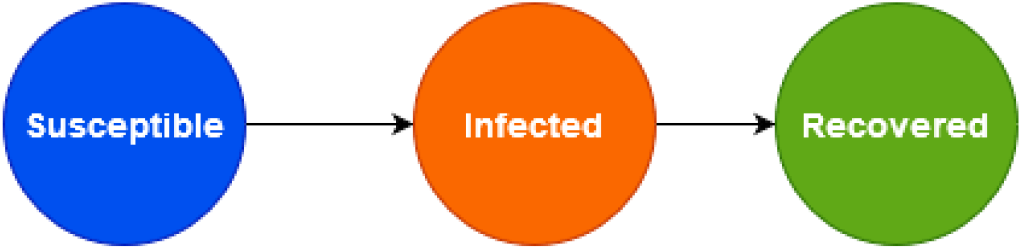
Population flow for SIR Model.

### 2.2 Equations

Three differential equations make the SIR Model. They are as follows:

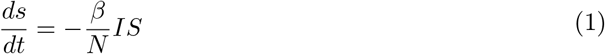

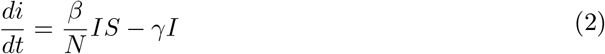

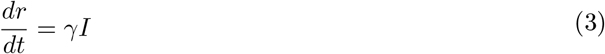

Where *t* is the time, *S*(*t*) represents the population susceptible to infection at *t, I = I*(*t*) represents the population infected at *t, R*(*t*) represents the population that has recovered at *t*. *β* denotes the contact rate, and 1/ *γ* denotes the average infectious period. N denotes total population size. From the above three equations, we get a total population size N governed by the equation:

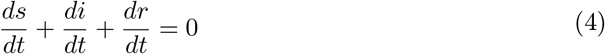

i.e. *N* = *S* + *I* + *R* = *constant*

For SIR Model, R_0_ (Basic Reproduction Number) is obtained from the following equation:

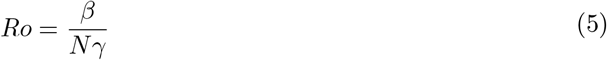

### 2.3 Forecasting of India

Considering the equations of the SIR model, Equations (1), (2) and (3) we try to simplify the assumption. Since the population in question is very large, the impact of the infection is insignificant relative to the size of the population. In a country of population 1.33 billion where the current number of deaths due to corona virus is 8102 [5], the percentage of infections is very small when compared to the population i.e approximately 0.00000609172. Therefore, Equation (2) becomes,

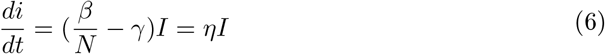

Where 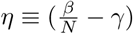

By integral calculus we have,

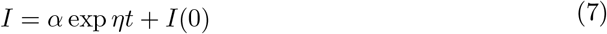

Thus, Equation (7) shows us that the growth of the epidemic will initially be exponential as in Figure 2.

**Fig. 2.**
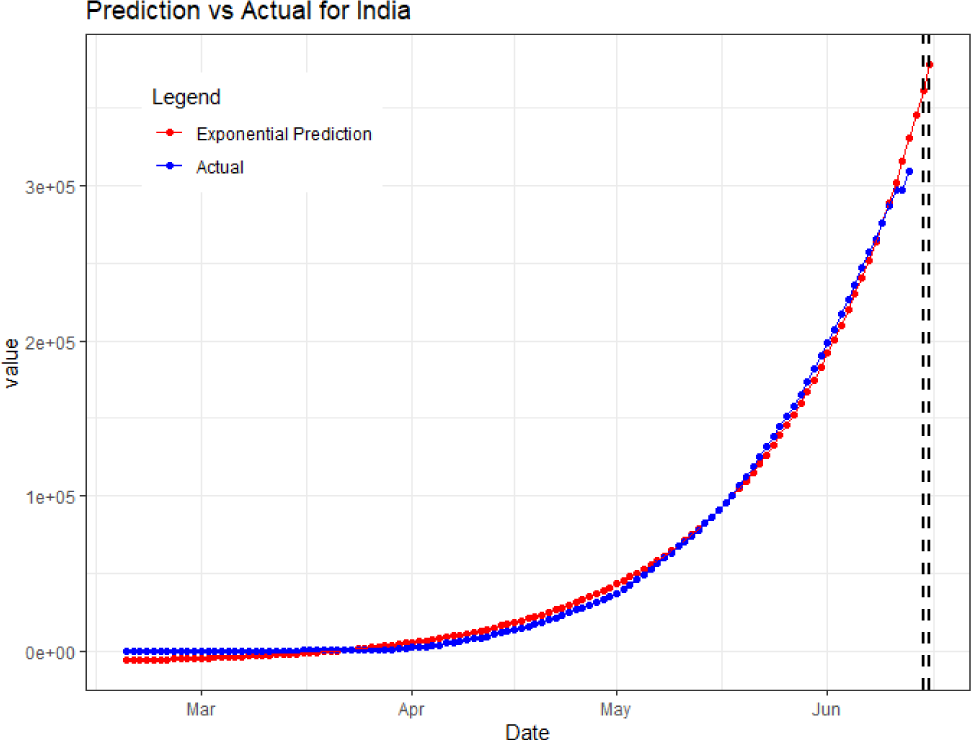
Diagrammatic view of Predicted vs Actual number of confirmed cases in India. Left vertical dashed line represents 12^*th*^ June 2020 and right vertical dashed line represents 13^*th*^ June 2020.

### 2.4 Forecasting for Iran

For forecasting the number of confirmed cases in Iran for 12^*th*^ June 2020 and 13^*th*^ June 2020, Equation (7) is used. Inspection ofFigure 3 reveals the exponential growth of the number of cases in Iran.

**Fig. 3.**
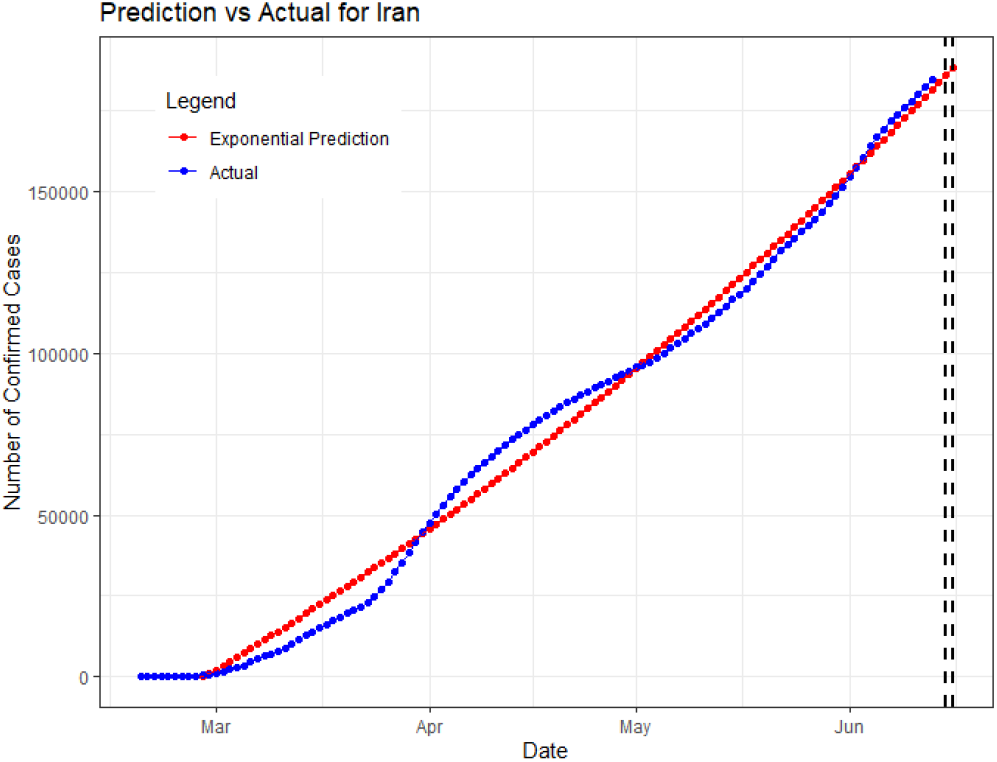
Diagrammatic view of Predicted vs Actual number of confirmed cases in Iran. Left vertical dashed line represents 12^*th*^ June 2020 and right vertical dashed line represents 13^*th*^ June 2020.

## 3 Discussion

A discussion of the different factors that influence a quarantine and the necessary precautions that must be taken for it are presented. We then delve into the measures put in place in China, Italy, Iran and India to determine how effective these strategies have been.

### 3.1 Factors and Precautions

The ability to anticipate the possible spread of the outbreak is important to devise strategies for effective resource allocation as well as implementing various intervention policies in favor of public health. For this, identification of appropriate factors is necessary. Consideration of population density, geographic land area, and spatial demographic data is critical so as to curb the growth of the outbreak. Considering the mobility of humans is essential in understanding the spread of the disease geographically but unfortunately, the data regarding the dynamics of mobility on spatial and temporal scales limit the accuracy of the forecast. Environmental and host susceptibility can help interpret the potential drivers of the infection and can improve the efficiency of forecasting and implementing policies of public interest [17]. A quarantine of 14 days must be taken by any person that comes in contact with a laboratory-confirmed case of infection. People in quarantine should be placed in spacious rooms with adequate ventilation facilities. Ensuring that people who are quarantined practice proper hand hygiene should be done. Environmental disinfection processes should be carried out consistently. Quarantined people should be examined daily [18].

### 3.2 China

On 31^*th*^ December, China reported cases of an unknown respiratory illness in Wuhan to WHO. The laboratory-confirmed cases in China on 2^*nd*^ January had reached 41. China started to work on identifying the novel virus and reported no new cases for the following 16 days. The number of cases rose to 62 in China on 20^*th*^ January. The government of Wuhan went ahead with the celebration of the Chinese New Year with 40,000 families on the very same day. The first case, outside Wuhan, was reported on 19^*th*^ January. The total number of cases increased to 548 on 22^*nd*^ January. During the first 20 days of January, very few quarantine measures were introduced and this led to more than 500 in less than 20 days. China did not act as strictly as was demanded by COVID-19 during its early stages. Wuhan (population : 11 million) being one of the most populous cities of China, did not help.

Quarantine measures were introduced in Wuhan on 23^*rd*^ January 2020. Public transportation and outward bound flights and trains were suspended. The government of Wuhan advised citizens to use masks. Zhejiang, Guangdong, and Hunan announced the highest level of public health emergency; level 1. Various hospitals were assigned to receive patients suffering from fever of unknown cause. Wuhan began construction of spe-cial emergency hospitals to combat COVID-19. Every province in China had declared a level 1 public health emergency. Various tourist attractions and cinema halls were also closed. By 26^*th*^January 2020, China began nationwide monitoring at airports, railway stations, bus stations and ports for testing and isolation of anyone infected. Wuhan suspended visa and passport services for Chinese citizens until 30^*th*^ January. Interprovincial transport was halted in some provinces. The percentage increase in cases was on a rise for the last 10 days of January (see Figure 4) due to the lack of quarantine measures during the early outbreak of the disease. Another reason could be the smaller measures that could have been implemented. For example, Schools in Beijing were closed until further notice on 26^*th*^ January 2020. But Beijing still wasn’t on lock down. By the end of January various malls were shut all over China, but some malls in Wuhan, the epicenter of the outbreak, were still operating for shorter periods of time. At the end of January, the number of cases in China were almost 10,000, most of them from Wuhan, Hubei with 200 deaths. This highlights the fact that China should have introduced quarantine measures sooner, not only to control its spread within China, but also to other countries.

**Fig. 4.**
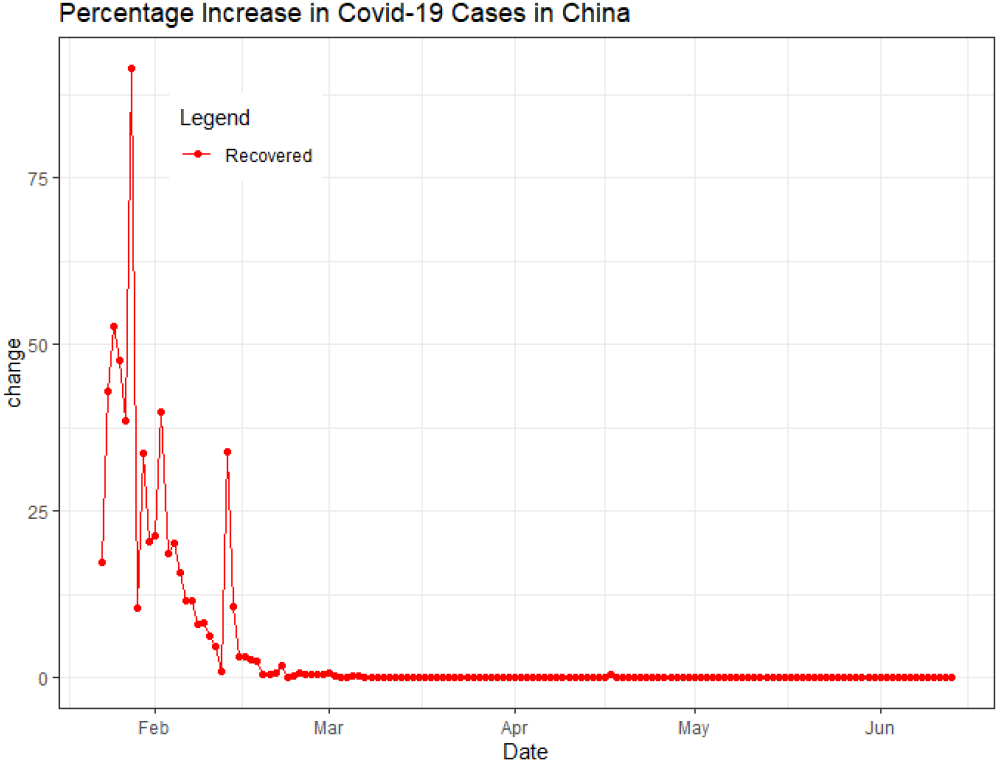
Plot of the percentage rate of change in the confirmed cases in China from 23^*rd*^ January 2020 to 13^*th*^ June 2020.

February was marked by a decrease in the rate at which individuals were being infected and an increase in the recovering population Figure 5. To contain COVID-19, Hubei implemented a stern policy which only allowed shop workers and one individual per household to be outside, medical reasons being an exception. After a patient’s feces tested positive for COVID-19 in Shen-zhen, China also took measures to prevent the spread of COVID-19 through sewage. On 3^*rd*^ February 2020, the Government of Wuhan declared that all the patients who had been in any contact with an infected person should be quarantined. By 5^*th*^ February, shelter hospitals were up and running in Wuhan for taking in the patients who weren’t suspected or confirmed. To prompt citizens to go see a doctor in case of fever or cough, some provinces temporarily banned the sale of fever and cough medicines in retail pharmacies. Hubei didn’t allow the sale of epidemic-related items at more than 15% of the stocking price. Even though there was a decline in the rate of increase of infections, the number of new cases were increasing. The graph in Figure 4 starts becoming flat somewhere around 15^*th*^ February 2020. Wuhan went on complete lock down on 13^*th*^ February. By 16^*th*^ February, no one was allowed to be out on the streets except for epidemic-related reasons and public places were closed. Not following any of these policies had various repercussions. The effect of these measures can be seen in Fig as by 20^*th*^ February, the percentage increase in the number of infections in China was below 1%. Within the next 2-3 days, various provinces relaxed their public health emergency levels from level 1 to level 2 and 3. Wuhan relaxed its quarantine measures by allowing people to leave the city on certain conditions with continuous monitoring. People of Wuhan were still asked to take their temperature twice a day and report if it was higher than 99.1F. Those who had recovered from COVID-19 were put on a 14-day quarantine. By the end of February, various provinces began screening people coming from countries like Japan, South Korea etc. In March, the number of new cases saw a huge drop and on 2^*nd*^ March 2020, Wuhan closed its first hospital that was constructed for the epidemic. Wuhan closed 10 more of its COVID-19 hospitals. Their effective quarantine measures in the month of February brought down their percentage increase in new cases to less than 0.1%. The current scenario is such that there are less than 10 new cases every day and less than 100 active cases.

**Fig. 5.**
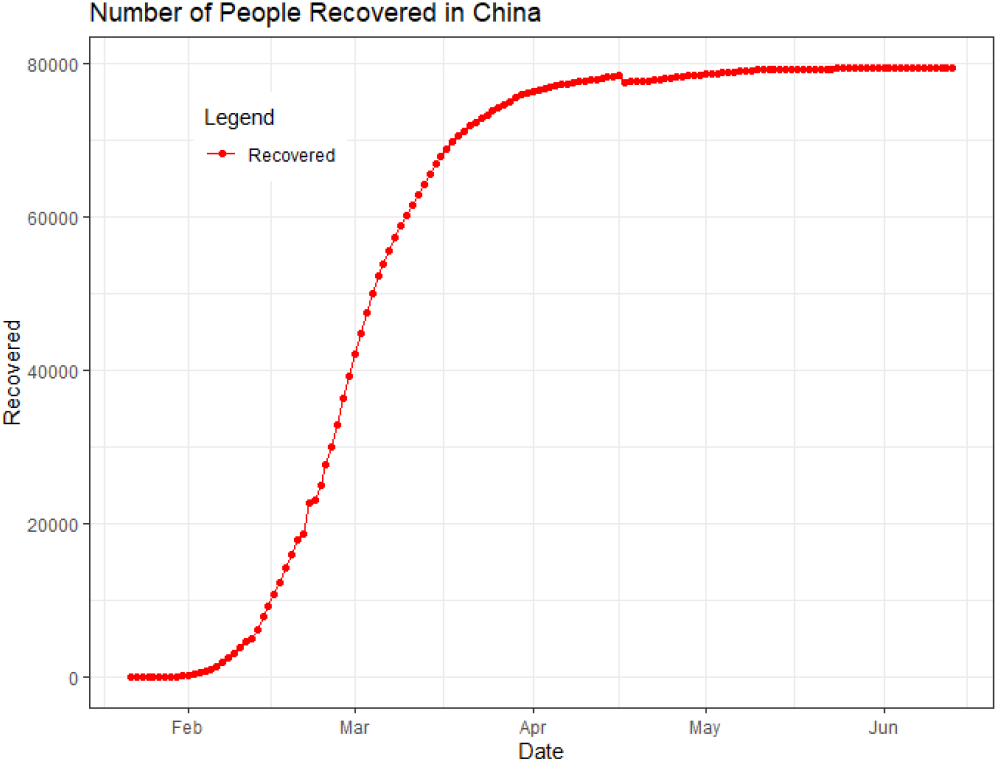
Representation of the number of recovered cases recorded in China rate from 23^*rd*^ January 2020 to 13^*th*^June 2020.

### 3.3 Italy

The first cases of COVID-19 were confirmed in Italy on 31^*st*^ January, travelers from Wuhan. They arrived in Italy on 23^*rd*^ January 2020 and tested positive on 31^*st*^ January 2020. The government introduced thermal screening and temperature checks at airports for international passengers. After a lull of almost 20 days, 16 new cases were confirmed in Italy. Most of which were from Lombardy, where no one had tested positive earlier. The government of Italy didn’t take strict precautions during the first 20 days of February. But, as the number of cases more than tripled from 21st February to 22^*nd*^ February (20 to 62), they declared quarantine in some provinces of Northern Italy, covering around 50,000 people. Quarantine zones, called red zones, prohibited people from traveling except for medical reasons. These red zones had various checkpoints to monitor the entry and exit of people. But there were multiple cases of people easily escaping these red zones. Other parts of Italy quarantined people traveling from Northern Italy for 14-days.

By 24^*th*^ February, 4 out of every 5 cases experienced mild or no symptoms. This came shortly after the surge in the number of cases from 22^*nd*^ February. By 25^*th*^ February, schools and colleges were closed and various companies instituted work from home. The number of cases became 1128 on 29^*th*^ February. The major reason for this was that initially, only patients coming from red zones were tested. But once all patients with symptoms were tested the number of cases in Italy increased. The more crucial period, was when asymptomatic-people didn’t take necessary precautions and transmitted the disease unknowingly. On 1^*st*^ March Italy segmented its territory into red zones, yellow zones and the remaining parts implemented safety and hygiene regulations i.e., left public places open. Red zones quarantined the entire population, whereas yellow zones closed schools and cinema halls etc. As the number of deaths crossed 100, on 4^*th*^ March, Italy closed schools and universities until 15^*th*^ March, with crowd control measures instituted. On 8^*th*^ March, more than 16 million people were placed under quarantine in Lombardy. Additionally, there was closure of several venues across the country. With the amount of cases at almost 10,000 and deaths more than 400, Italy went on complete lock down on 9^*th*^ March. The government restricted non-essential travel. As a result, swarms of people rushed to the supermarkets and prison riots erupted. On March 11, it was announced by the Prime Minister to shut down all stores except grocery and pharmacy shops till 25^*th*^ March. Closure of eateries and bars but not of public transportation or postal services was announced. In Italy, the number of infected people and deaths were increasing at an alarming rate Figure 6. This can be attributed to the fact that Italy is marked with the second largest percentage of old people globally with as many as 28% of Italians falling in the age group of 60 and above. COVID-19 is known to be more lethal for older people due to their weaker and compromised immune systems. But the rate of increase of reported cases in Italy was worrisome. It highlighted the measures that should have been taken by the Italian government such as conducting enough tests during the early stages to contain COVID-19. Earlier detection of cases could have led to earlier isolation. Additionally, the red zones in Italy were not well-managed, further spreading the disease. The Italian government couldn’t prepare their medical resources. Many medical facilities were operating at full capacity and therefore were unable to treat each and every patient. If Italy had been more proactive with testing people in the early days of February, then the situation could have been a lot better.

**Fig. 6.**
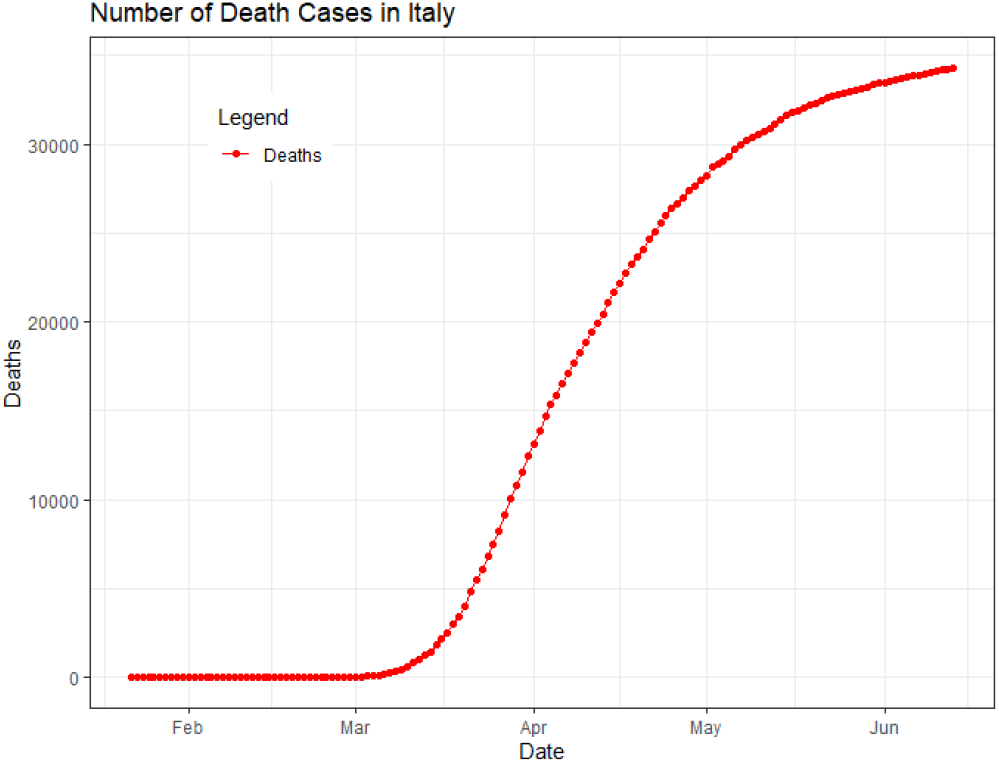
Representation of the number of cases of Deaths recorded in Italy from 23^*rd*^ January 2020 to 11^*th*^ June 2020.

### 3.4 Iran

Iran first reported a case of coronavirus on 19^*th*^ February, when two people had tested positive in Qom. Both were announced dead later in the day. Despite a letter requesting limitations on the number of pilgrims at the shrines in Qom, they remained open for congregation in fact, one of the heads of the shrines encouraged visit to it on 27^*th*^ February. It was only on 16^*th*^ March that this shrine along with a few others was closed. Protests followed outside the shrine and servants had to drive out the protesters.

By 22^*nd*^ February, all concerts and cultural/ sports events were canceled. The Universities and Educational institutions were also closed in various provinces. On 26^*th*^ February, the president announced their plans to Quarantine only the infected individuals and not entire provinces. Friday prayers were canceled in Tehran and other gravely infected regions. The number of reported cases had grown to 978 Figure 7 of which 23 were members of the Parliament. By the next day, (2^*nd*^ March) this number became 1501. On 3^*rd*^ March, Iran executed temporary release of over 54,000 prisoners to curb spreading of infections in crowded jails and announced plans involving as many as 300,000 soldiers/ volunteers. Several countries including, Singapore, India, New Zealand placed restrictions on travel to Iran. On 5^*th*^ March, 591 new cases were observed. This was followed by the parliament being suspended and international travel being banned for government officials. There was an announcement of plans for checkpoints between different cities to limit travel. Schools and universities were closed 28th March. According to a report on 7^*th*^ March, 1669 had recovered and 16000 had been hospitalized as suspected positives for testing purposes. That Iran was making facilities available for treatment in every province was reported by a WHO representative placed in Iran. On 9^*th*^ March, up to 70000 prisoners had been released temporarily. By 14^*th*^ March, cases had risen to 12,729 and deaths to 611. A statement was made by the supreme leader on 19^*th*^ March to forbid unessential travel. 85000 prisoners were released. The celebration of the traditional fire festival of Persians was forbidden. The economic sanctions make Iran’s health care system weaker and unable to deal with the outbreak. The public resentment towards containment measures and distrust of officials resulted in unsuccessful containment efforts. The clerics objected to a lock down and parts of the public were involved in acts of defiance.

**Fig. 7.**
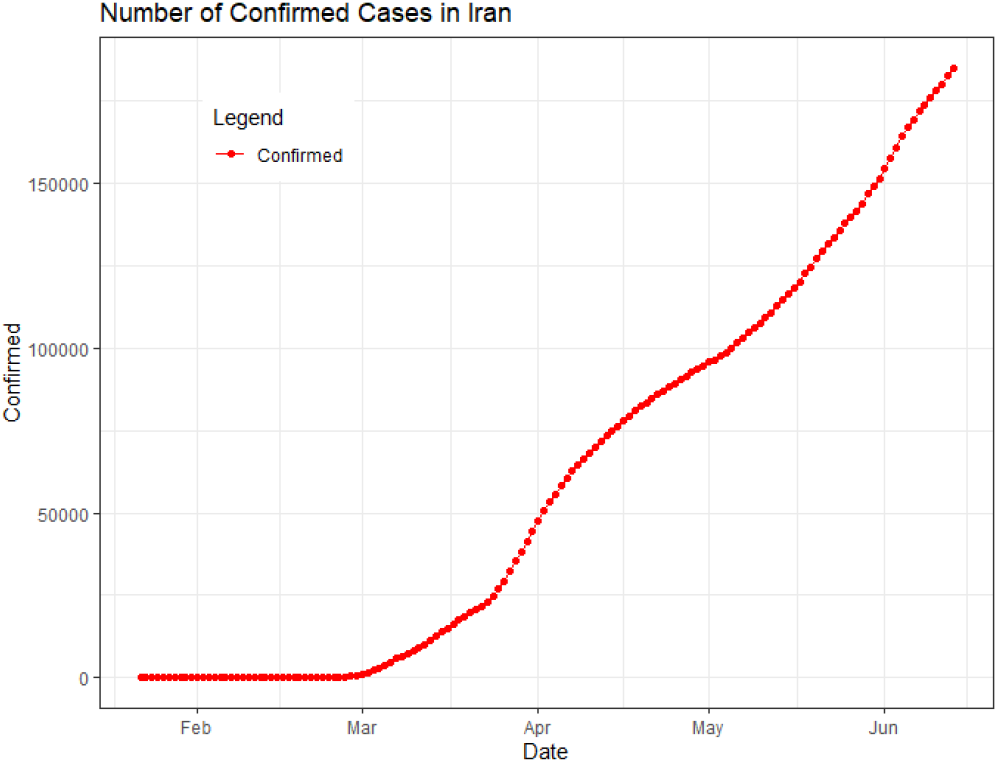
Representation of the number of cases in Iran from 23^*rd*^ January 2020 to 11^*th*^ June 2020.

### 3.5 India

As on the 14^*th*^ of June 2020, the number of COVID-19 cases in India had risen to 320,922 with as many as 149,348 active cases and 162,378 people recovered one. While the statistics report a higher number of recovered cases than active ones, the strategy and planning that brought about this result are worth examining. The mortality rate as observed thus far was 2.8 per cent.

The first case made an appearance on 30^*th*^ January 2020. However, at that point measures of airport screening had been implemented in 20 of the country’s airports. Although initially, only arrivals from China were screened, the list included several other countries by the end of February. The month of February did not show a significant increase in cases. As the first death due to COVID-19 occurred on 12^*th*^ March, all non-essential visas were canceled and worry began to set in among the population. The government responded by strongly advising the citizens to follow social distancing protocols. As the end of March drew closer, a nationwide lockdown was imposed on 24^*th*^ March to be followed for the duration of about three weeks. The number of cases had exceeded 1000 on 29^*th*^ March and doubled in the next 4 days. The lockdown ensured the closure of educational institutions and public facilities for a period of 3 weeks across the states and union territories where confirmed cases had been recorded. It was able to slow down the spread of the pandemic to a good extent. This can be inferred from the graph of confirmed cases in India Figure 2 as compared to that of Iran Figure 3. In India, this curve is seen to be of an increasing nature, however, the rate of increase of this slope is much lower than that of Iran. In Iran, the number of confirmed cases shot up very rapidly.

Regardless, the number of daily cases were still rising and predictably soFigure 8. They had risen to over a thousand on 13^*th*^ April and didn’t fall below 800 until the 21^*st*^ from when they have been only greater than a thousand climbing to peaks of 12,375 on June 10^*th*^. In an attempt to continue slowing the spread of the disease, the lockdown was extended: first on 14^*th*^ April for another three weeks and once again on 1^*st*^ May till 17^*th*^ May and finally, it was extended till 31^*st*^ May. It is worth noting that with each new lockdown being announced, the states where the spread had been contained to no new cases in a designated period of time, saw some relaxation in measures. The containment zones remained regions of strict quarantine rules but the free movement in the regions not labeled containment zones has become an alarming concern because they can still be conducive to community transmission. With an average population density more than 400 people per square km, the norms of social distancing are not realized when the lockdowns and curfews end.

**Fig. 8.**
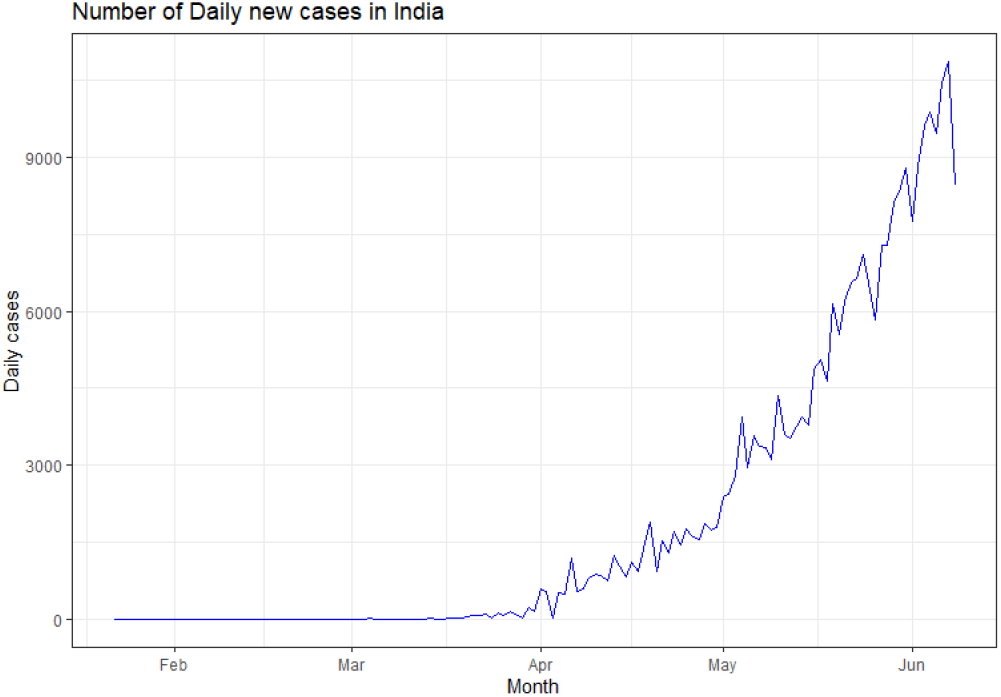
Line plot of the number of new cases daily cases in India from the month of February to June.

## 4 Results

The forecast values of confirmed cases for India and Iran are shown in Table 1 and Table 2 respectively.

**Table 1.** Predicted number of Confirmed Cases for India

| Status | 12<sup>th</sup> June 2020 | 13<sup>th</sup> June 2020 |
| --- | --- | --- |
| Actual | 297535 | 308993 |
| Predicted | 317669 | 332633 |

**Table 2.** Predicted number of Confirmed Cases Iran

| Status | 12<sup>th</sup> June 2020 | 13<sup>th</sup> June 2020 |
| --- | --- | --- |
| Actual | 182525 | 184955 |
| Predicted | 186735 | 189645 |

The Accuracy metric used is the ratio between the sum of difference between actual and predicted cases on 12^*th*^ June 2020 and 13^*th*^ June 2020 and the total number of actual cases on 12^*th*^ June 2020 and 13^*th*^ June 2020. The accuracy of forecasting the confirmed cases in India and Iran obtained is 92.78% and 97.57% respectively.

## 5 Conclusion

Although China Implemented quarantine measures a little late, it made sure to correct the fallout by imposing strict regulations and taking the necessary steps. The release of medical resources as required reduced the infected population of China to a huge extent. The epicenter of the outbreak shifted to Italy with Iran becoming the third most affected country as declared by WHO soon after. From the management of medical resources in Italy, one can infer that this outbreak must be taken seriously with more emphasis on precaution than cure. Iran became a contributor in spreading the disease to other countries as it did not have any strict regulations on international travel. India rightly placed travel restrictions on all international flights. While the Indian government was able to slow down the spread to a significant extent in the early stages of the lockdown, the statistics of the months of May show a rise in the number of infected. This can be attributed to the relaxation both on the account of the government and the one self-imposed by the citizens who have been cramped at home for the past two months. It is this attitude of despair and muted exasperation coupled with the high percentage of recovery cases that creates a false sense of confidence in the public towards being able to fight off the pandemic. But if the statistics show us anything and if the saturated and exhausted healthcare professionals, government representatives and policemen and policewomen are evidence of anything, it is that taking the pandemic lightly is not something the country can afford.

## Data Availability

All data used in the research paper is available publicly.

https://github.com/CSSEGISandData/COVID-19/tree/master/csse_covid_19_data/csse_covid_19_time_series

## 6 Data Availabilty

The data set made use of in this paper is from WHO. It is also used by JHU in the interactive Map on their website. It is available to the public. This data set encompasses the cases of Confirmed, Recovered and Deaths associated with coronavirus outbreak in as many as 157 countries/regions at the time of this research. The data used in this research spanned from the very beginning of cases in China as on 22^*nd*^ January 2020 up to 13^*th*^ June 2020.

